# High definition analyses of single cohort, whole genome sequencing data provides a direct route to defining sub-phenotypes and personalising medicine

**DOI:** 10.1101/2021.08.28.21262560

**Authors:** KE Joyce, E Onabanjo, S Brownlow, F Nur, KO Olupona, K Fakayode, M Sroya, G Thomas, T Ferguson, J Redhead, CM Millar, N Cooper, DM Layton, F Boardman-Pretty, MJ Caulfield, Genomics England Research Consortium, CL Shovlin

## Abstract

Possession of a clinical or molecular disease label alters the context in which life-course events operate, but rarely explains the phenotypic variability observed by clinicians. Whole genome sequencing of unselected endothelial vasculopathy patients demonstrated more than a third had rare, likely deleterious variants in clinically-relevant genes unrelated to their vasculopathy (1 in 10 within platelet genes; 1 in 8 within coagulation genes; and 1 in 4 within erythrocyte hemolytic genes). High erythrocyte membrane variant rates paralleled genomic damage and prevalence indices in the general population. In blinded analyses, patients with greater hemorrhagic severity that had been attributed solely to their vasculopathy had more deleterious variants in platelet (Spearman ρ=0.25, p=0.008) and coagulation (Spearman ρ=0.21, p=0.024) genes. We conclude that rare diseases can provide insights for medicine beyond their primary pathophysiology, and propose a framework based on rare variants to inform interpretative approaches to accelerate clinical impact from whole genome sequencing.

## INTRODUCTION

High definition medicine offers opportunities to advance clinical care from single disease-based approaches, where trials of thousands of individuals are used to identify best practice, to a tailored approach for each patient encompassing multi-morbidities and health enhancers. Personalising medicine is anticipated not only to enhance efficacy and efficiency of interventions, but also to reduce adverse events from unnecessary investigations or treatments.

Sequencing the whole genome provides one potential blueprint for such personalisation, though the challenges required for full potential to be realised cannot be over-estimated.[1] Furthermore, delivering numeric and clinical-level confidence to guide personalised medicine is demanding in the setting of individually unique genomes. We hypothesised that some challenges could be overcome in a generally instructive manner by applying whole genome sequencing to a set of well characterised endothelial vasculopathies where the abnormal vascular structures may unmask deleterious potential of variants in disease-independent genes that would usually result in subclinical hematological phenotypes.

The most common of the endothelial vasculopathies, hereditary hemorrhagic telangiectasia (HHT), has established clinical diagnostic criteria, known causal genes, and expert consensus informing clinical practice across general and sub-speciality management.[2-4] Estimated to affect approximately 1.5 million individuals worldwide,[5,6] HHT usually results from a heterozygous loss-of-function allele in *ENG, ACVRL1* or *SMAD4*, and is inherited as an autosomal dominant trait.[7] Haploinsufficiency at one of these loci predisposes to the development of abnormal vascular structures, especially arteriovenous malformations (AVMs) characterised by turbulent flow and defective vascular walls predisposing to hemorrhage.[8] Affected individuals experience spontaneous, recurrent nosebleeds due to abnormal nasal vasculature, and exhibit small visible telangiectatic vessels that tend to develop on the lips, oral cavity, and finger pads.[3,4] Many have long-term excellent health with modest hemorrhagic losses, though most experience nosebleeds sufficient to lead to iron deficiency anemia unless dietary iron intake is supplemented,[9] and significant proportions require surgical interventional treatments, and/or disease modifying drugs such as anti-angiogenic agents.[2,3,10] Patients are commonly found to have silent visceral AVMs, where screening/primary prevention strategies have led to improved life expectancy.[11,12]

As is typical in medicine, it remains impossible to predict which patients will develop complications prior to, and/or despite preventative treatments. Marked diversity in vascular malformations between affected members of the same family were emphasised in the earliest genetic studies,[13] but more strikingly, similar vascular abnormalities can have profoundly different impacts: HHT hemorrhagic severity varies both in severity of recurrent bleeds from nasal or gastrointestinal telangiectasia,[2-4] and rare but life-threatening acute hemorrhages from pulmonary or cerebral AVMs.[14,15] Inflammation is not a prominent feature of HHT,[9] and the limited available data suggest iron handling is appropriate,[9] but in one study, anemia was out-of-proportion to hemorrhagic iron losses in more than a quarter of consecutive hospital-reviewed patients, with evidence for reduced red blood cell (erythrocyte) survival in nearly half of these.[16] Counterintuitively, age-adjusted incidence rates of venous thromboembolism (VTE) are several-fold higher in HHT than in the general population,[17,18] while HHT-affected individuals who have VTE are more likely to have a major deep-seated infection that is a particular risk when venous blood can evade pulmonary capillary processing by passage through right-to-left shunts provided by pulmonary AVMs.[19-22] Socio-environmental exposures can aggravate HHT, as detailed elsewhere,[3,16,17,20,23-30] but only a proportion of patients with these added “risk factors” develop the respective clinical phenotypes. There are hints of familial predispositions [16,17,20,23,24,27] supported by case series where occasional individuals had coincidental non-HHT genetic contributors to their complications.[16,17]

Here we report the magnitude of deleterious genomic variability across clinically-relevant genes, categorised to enhance statistical power, in an essentially unselected cohort of vasculopathy patients who had undergone whole genome sequencing through the National Health Service’s 100,000 Genomes Project.

## RESULTS

### Patient cohort demographics

104 individuals were recruited with HHT from a single Genomic Medicine Centre. They were from 89 families. At the time of recruitment, patient ages ranged from 17-87 years (median 50 years), and 66 (64%) were female. 56/89 (63%) of families were found to have heterozygous variants in one of the 6 genes currently on HHT (endothelial vasculopathy) gene testing panels in the UK National Health Service, namely *ENG, ACVRL1, SMAD4, GDF2, RASA1*, and *EPHB4* (*Figure 1A*). In keeping with the “HHT gene negative” status of a proportion of families on recruitment, in 33/89 (37%) of families, no variants were identified in known HHT genes.

**Figure 1:**
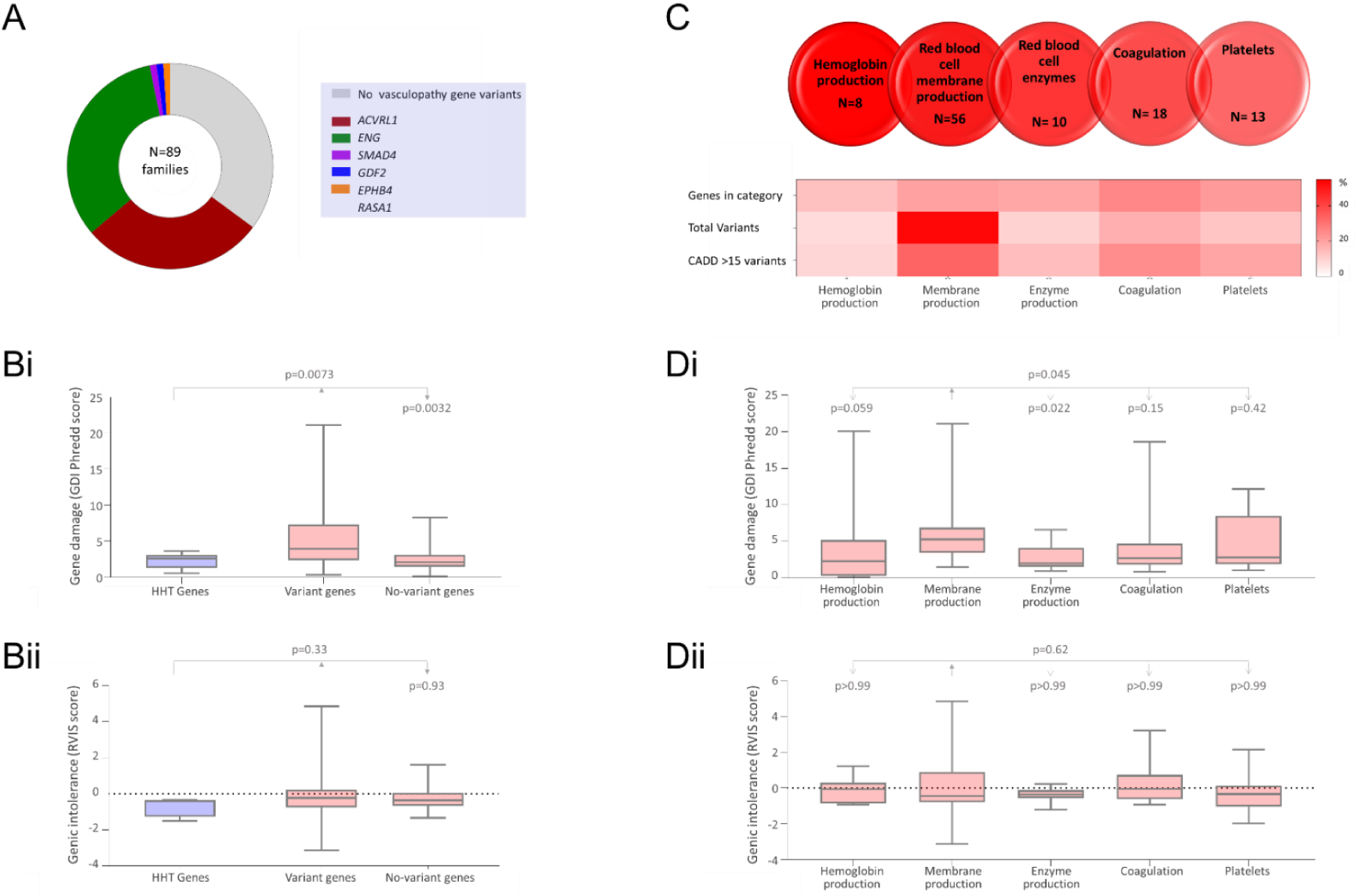
Gene-level metrics. **A)** Endothelial vasculopathy genes and variants identified in current study cohort. Note the major HHT genes are *ENG* and *ACVRL1*,[7] while *EPHB4* and *RASA1* cause separate endothelial vasculopathies (CM-AVM2 and CM-AVM1) which overlap phenotypically with HHT. **B)** Population-level burden of genetic damage in the 6 endothelial vasculopathy genes (blue), and 75 study hematological genes (red), as detailed in *Supplementary Table 1*. The study genes were sub-categorised by variant presence (‘variant genes’, N=56) and absence (‘no-variant genes’, N=19) in the study cohort. **Bi)** Gene damage index (GDI) Phred scores.[31] **Bii)** Residual variation intolerance scores (RVIS).[32] **C)** The five process-level categories: Respective category contributions in the study cohort are shown for the total number of genes, variants, and variants with Combined Annotation Depletion (CADD[35-37]) score >15 (likely deleterious, as described further in text): Each box represents the percentage for the row, as indicated by the heat scale. **D)** Variation burden within the general population for the 75 study genes in the respective categories: **Di)** Gene damage index (GDI) Phred scores [31], and **Dii)** Residual variation intolerance scores (RVIS).[32] All box plots indicate median and interquartile range, error bars full range. Overall p values were calculated by Kruskal Wallis with Dunn’s multiple comparison test used to derive p-values for pre-selected pair-wise comparisons indicated by arrows.

### Gene-level variation

A total of 105 variants were identified within the cohort in the 75 selected hematological genes (*Supplementary Table 1*). At least one variant was present in 52/104 (50%) of the study cohort, and 56/75 (75%) of genes had at least one variant identified in the study cohort.

The variant burden was not specific to the study cohort, as the gene damage index (GDI [31]) was also greater in the genes harboring variants in the study cohort (*Figure 1Bi*), providing evidence that these genes are more damaged in the general population. There was no difference between the variant and non-variant genes for residual variation intolerance (RVIS, *Figure 1Bii*), a measure of genic intolerance that performs better for detecting causality for recognised diseases.[32]

The variants were identified in all five categories of hematological genes, i.e. in the major categories of congenital hemolytic anemias (hemoglobin, erythrocyte membrane and erythrocyte enzyme production),[33] together with coagulation and platelet genes relevant to hemorrhagic and thrombotic states [34] (*Figure 1Ci*). Variants were over-represented in the red cell membrane production category although the excess was less pronounced for variants predicted to be more deleterious (*Figure 1Ci*). It was noted that population-wide differences between the five categories of genes primarily reflected lower gene damage indices in the enzymopathy and hemoglobin categories (*Figure 1D*).

Variants were identified in genes on all chromosomes except Chr21 and ChrY (*Figure 2Ai*). The number of variants per Mb of selected gene loci varied in accordance with locus positions for genes with high GDI and RVIS scores (*Figure 2Ai*). Variant-containing genes were also distributed throughout the genome for patients with variants in the HHT genes *ACVRL1* and *ENG* (*Figure 2Aii*). All variants were rare in the general population, with GnomAD 3.1.1 [38] allele frequencies less than 0.3%, median GnomAD allele frequency less than 0.05% for each of the five categories, and no significant differences between the 5 categories of genes (Kruskal Wallis p=0.53, *Figure 2B*).

**Figure 2:**
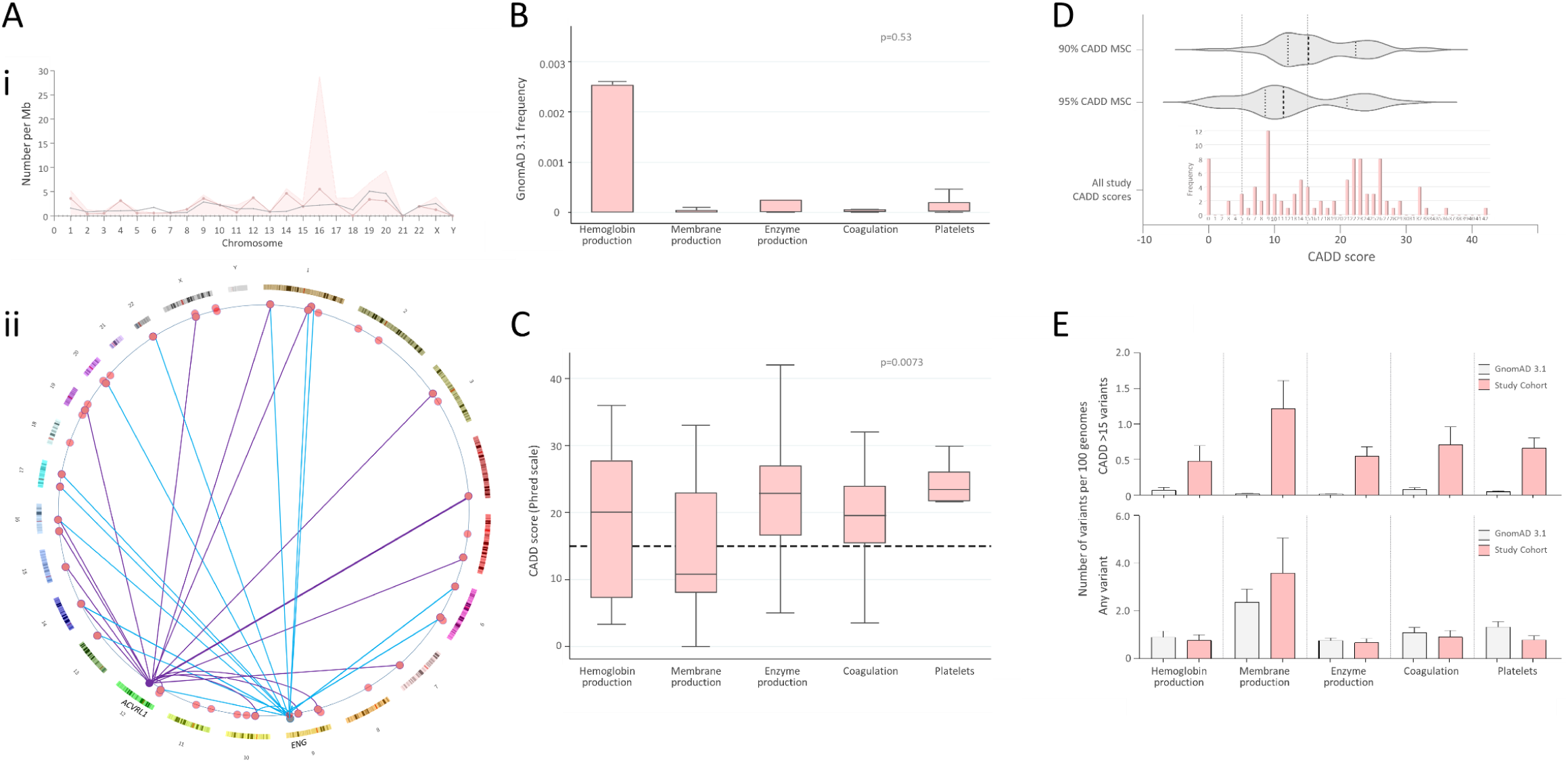
Variant-level metrics. **A)** Genome positions of the 56 variant-containing genes in the study cohort. **Ai)** Chromosomal distributions of variant-containing genes per 100Mb of DNA (grey circles/lines), number of variants per Mb (red shaded background), and variants with CADD score >15 (red circles/lines), per Mb of gDNA by chromosome [39]. **Aii)** Genome ideogram [40,41] indicating positions of the 56 study genes with variants (red), and the two major HHT genes *ACVRL1* and *ENG* (grey). Variants identified in patients with an identified variant in HHT genes are indicated for *ACVRL1* (purple lines) and *ENG* (blue lines). To preserve anonymity, data for the single *SMAD4, GDF2* and *EPHB4* families are not illustrated. **B)** Allele frequencies in GnomAD 3.1.1,[38] and **C)** CADD scores [35-37] of all variants by category. Box plots indicate median and interquartile range, error bars 95% confidence intervals, with dots indicating outliers, horizontal line indicating CADD score of 15, and p-values calculated by Kruskal Wallis. **D)** Gene-level mutation significance cutoff (MSC) scores[42], indicating the lower limit of the 90^th^ and 95% confidence intervals for deleterious CADD scores in individual genes (grey violin plots) compared to the CADD score distribution of variants identified in the study cohort (red). Vertical dotted lines represent overall CADD scores 5 and 15, and the violin plot median and interquartile ranges. **E)** Number of variants in the 75 study genes by category, within the GnomAD 3.1 dataset (grey), and current cohort (red), plotted as number per 100 genomes. Lower graph, rate of any variants/100 genomes compared to the number/100genomes of likely pathogenic and loss of function variants in the same genes in GnomAD 3.1 dataset. Upper graph, rate of deleterious variants/100 genomes compared between the two cohorts-study cohort genes CADD score >15, GnomAD 3.1 as defined in Methods section.

### Variant deleteriousness

Variant CADD scores ranged from 1-42 (median 15), with the distribution differing between categories (p=0.0073, *Figure 2C*). Fifty-seven of 105 variants (54.3%) had CADD scores greater than 15, and at least one variant with a CADD score >15 was present in 38/104 (36.5%) of the study cohort.

Variants in the hemoglobin and red cell membrane production categories were more likely to have lower CADD scores than variants in enzyme, coagulation and platelet genes where more than 75% of CADD scores exceeded 15 (Kruskal Wallis p=0.0073, *Figure 2C*). Across all genes with variants, the CADD score delimiter of 15 captured the median 90% mutation significance cutoff (MSC) threshold (*Figure 2D*), while the CADD score delimiter of 5 captured the 90% MSC threshold of all except one gene (*Figure 2D*). In view of these findings, the simple categories of CADD score <5 (likely benign), 5-15 (uncertain significance) and >15 (likely deleterious) were retained generally for the purposes of the manuscript, but two CADD <15 variants in the outlying gene *SPTA1*, were tested in both uncertain significance and deleterious categories.

### Variant enrichment metrics

Gene damage indices and genic intolerance scores had suggested that there may be enrichment of variants in several categories within the study cohort (*Figure 1*). This was observed in the general population captured in GnomAD 3.1,[38] which, as in the study population, also contained a greater proportion of variants in red cell membrane genes (*Figure 2D*, lower panel).

A different pattern emerged for deleterious variants. All gene categories displayed at least 7.2-times more deleterious variants in the study cohort compared to GnomAD 3.1, per 100 genomes, in part reflecting the more liberal CADD score method to assign deleterious variants. However, enrichment varied between the categories, with a mean enrichment of 26-fold, and maximum 64-fold enrichment (*Figure 2D*, upper panel). By two-way ANOVA across all 75 genes, the 5 categories only accounted for 2.5% of the total variance (p=0.35), compared to 19.9% from the cohorts (p<0.0001). However, restricting to the 30 genes where deleterious variants were defined in both cohorts, while all variants were only enriched 1.2-2.4 fold (Kruskal Wallis p=0.63), deleterious variants were enriched 38.4 to 385-fold (Kruskal Wallis p=0.039).

### Patient subcategory classifications

To examine if presence or enrichment of deleterious variants may be biologically relevant in the study cohort, the variants were examined in the context of patient phenotypes. Blinded to the variants identified in hematological genes, the recruited patients had been assigned to relevant scales for more extreme phenotypes (*Figure 3A*). Notably, in this group of patients ‘selected’ purely due to the absence of a known family HHT genotype and attendance at the UK reference centre during recruitment to the 100,000 Genomes Project, 36/104 (35%) were categorised with severe hemorrhage, 29/104 (28%) with disproportionate anemia, and 8 (8%) had venous thromboembolic disease. Deep-seated infections affected 13/104 (13%): 10 were in association with concurrent and presumed causative pulmonary AVMs (cerebral and spinal abscesses, spinal discitis, and septic arthritis, due to polymicrobial flora, particularly anaerobic and aerobic commensals of the gastrointestinal and periodontal spaces, as described [19-21]).

**Figure 3:**
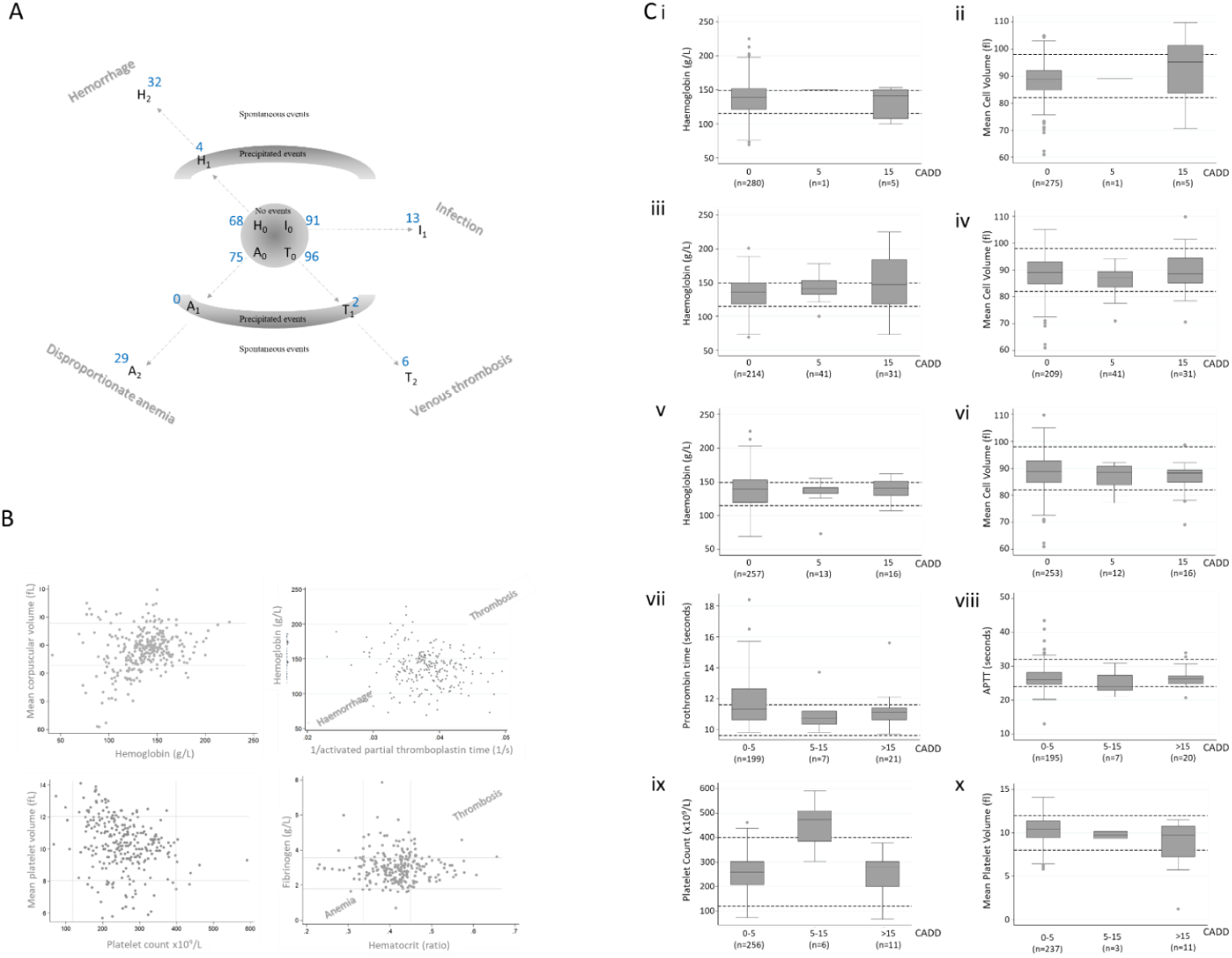
HHT patient subcategories and variation. **A)** Schematic of cohort of 104 plotted on 4 separate axes for hemorrhage [H], anemia [A], thrombosis [T], and deep seated infection [I]. Blue numbers indicate the number of the cohort with no events/no excess (Ho, Ao, To and Io), precipitated events (H1, A1, T1), and spontaneous events (H2, A2, T2 and I1). I1 were cerebral abscess (N=6), spinal discitis (N=2), spinal abscess (N=1), septic arthritis (N=1), recurrent bacterial endocarditis (N=1), osteomyelitis (N=1), and recurrent sepsis (N=1). **B)** Two-way distribution plots of all captured same-day measurements of hemoglobin, mean corpuscular volume (MCV), platelet counts, mean platelet volume (MPV), haematocrit, fibrinogen, and activated partial thromboplastin (APTT), with normal ranges indicated for MCV, MPV, platelet count, and fibrinogen. **C)** Quantitative phenotypic measurements categorised by presence, absence and CADD score of variants in **i/ii)** hemoglobin genes; **iii/iv)** red cell membrane genes; **v/vi)** red cell enzyme genes; **vii/viii)** coagulation genes; **ix/x)** platelet genes. Data points represent all datasets captured in the 104 patients across their clinical assessments. Two indices are shown per category, but there were no differences observed between categories for other erythrocyte indices (data not shown).

### Quantitative phenotypic traits

Quantitative traits differed between patients (*Figure 3B*). For each of the five categories of genes, the distribution of quantitative traits was compared between patients with no variants or CADD <5 variants (“likely benign”); CADD 5-15 variants (“uncertain significance), and CADD>15 variants (“likely deleterious”, *Figure 3C*).

For hemoglobin (*Figure 3Ci,ii*), red cell membrane (*Figure 3Ciii,iv*), and red cell enzyme (*Figure 3Cv,vi*) genes, there was no discernible difference in any measured red cell index by the presence of variants, with the median of each category within the respective normal range. Similarly, for coagulation genes, there was no discernible difference in prothrombin time or activated partial thromboplastin (APTT) time according to the presence of a variant (*Figure 3Cvii, viii*). The only category where a median value lay outside of the normal range was for platelet gene variants: platelet counts were higher for individuals who had variants with CADD scores between 5-15 (*Figure 3Cix*). Mean platelet volume did not differ where measured in the 3 of these (*Figure 3Cx*), and similar platelet count trends were not observed for the higher impact variants (*Figure 3Cix*). We concluded that the variants were not clearly reflected in variability within patient quantitative traits at specifically sampled times.

### Phenotypic categories

Next we examined gene variants across the categories by clinical phenotypes over the patients’ life-times to date. We restricted analyses to the presence of variants where CADD scores were >15, i.e. the ‘likely deleterious’ variant group. Variants with CADD scores >15 were present in all severe hemorrhage, disproportionate anemia, spontaneous thrombosis and severe deep-seated infection categories, but notably were also present in each of the 4 “no event” categories (*Figure 4A*).

**Figure 4:**
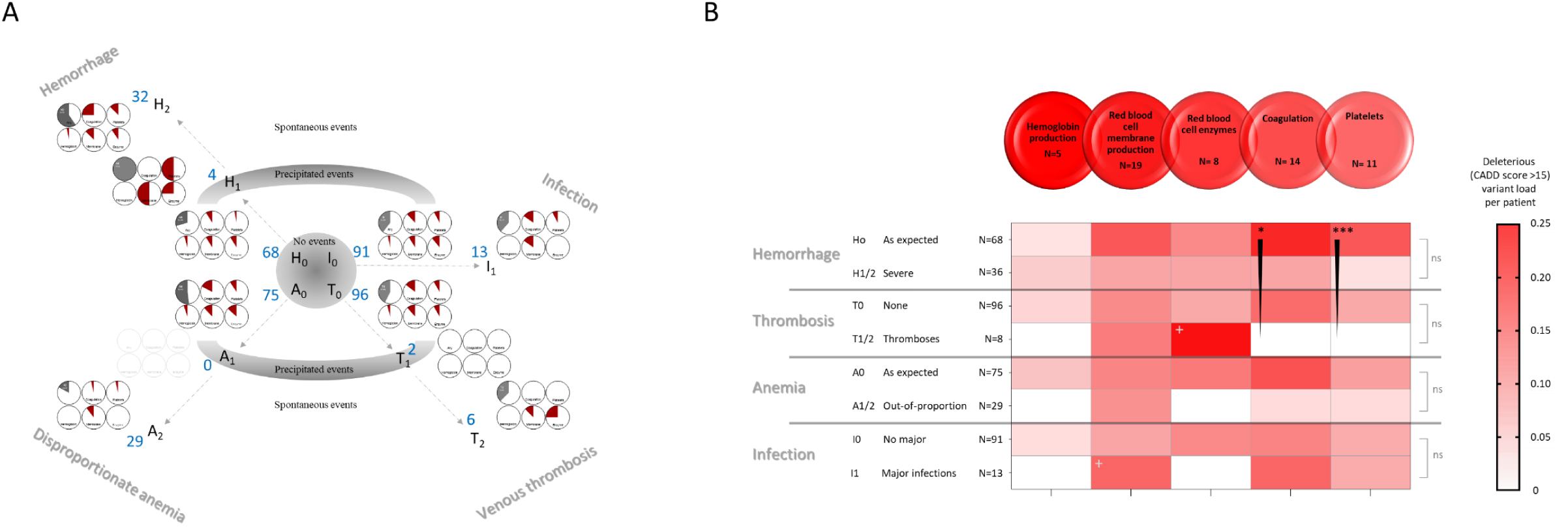
Burden of deleterious variants by categories of genes and phenotypic severity profiles. **A)** The burden of deleterious (CADD >15) variants in the 5 categories of genes for each pre-defined phenotypic subcohort. Filled pie-chart regions quantify the number of CADD>15 variants per patient in the category (grey if pooling across all gene categories, red if gene category-specific, fully filled if one variant per patient). The 10 variants with CADD scores of 1-5 (“likely benign”), and 36 with CADD scores between 5 and 15 (“of uncertain significance”) are effectively included in the remaining white pie chart regions, to enhance clarity. For each set of 6 pie charts, gene category placements are consistent. The upper row indicates total number of CADD>15 variants across all categories, and the remaining upper row pie charts focus on hemorrhage (upper middle for coagulation genes, upper right for platelet genes). The lower row presents the red cell gene categories (lower left for hemoglobin genes, lower middle for erythrocyte (red cell) membrane genes, and lower right for erythrocyte enzyme genes). The set of 6 grey-lined empty pie charts indicate there were no patients in the pre-defined A1 category. **B)** Heat maps for number of variants per patient in each phenotypic subcohort, where the precipitated and spontaneous H1/H2, T1/T2, and A1/A2 subcohorts have been pooled. Trends highlighted in the text are denoted by a grey cross, or black star where * indicates p<0.05, and *** p<0.01. The gene where MSC CADD score adjustments were made (*SPTA1*) was in the membrane category, and adjustment by MSC CADD did not materially alter these findings.

Hemoglobin gene variants with CADD scores >15 tended to distribute towards the “no event” categories (*Figure 4A*, lower left pie charts). Examining by binary categories that pooled precipitated and spontaneous events, deleterious hemoglobin gene variants were not enriched in any of the 4 phenotypic severity categories (*Figure 4B*).

Visual inspection suggested that erythrocyte membrane gene variants might be enriched in the 11 patients with severe deep-seated infections (*Figure 4A lower middle pie charts, Figure 5B*). There was no material difference denoting the *SPTA1* variants with CADD scores of 12.73 and 13.38 as deleterious (tested because *SPTA1* was an outlier gene with a 90% CADD MSC of 0.7), since one individual already had another variant in the category with a CADD score >15, and the second individual did not have a deep-seated infection. Similarly, visual inspection suggested that erythrocyte enzyme variants might be enriched in the 6 patients with unprovoked venous thromboemboli (*Figure 4A lower right pie charts, Figure 5B*). However, these were based on small numbers, and unsuited to statistical examination.

**Figure 5.**
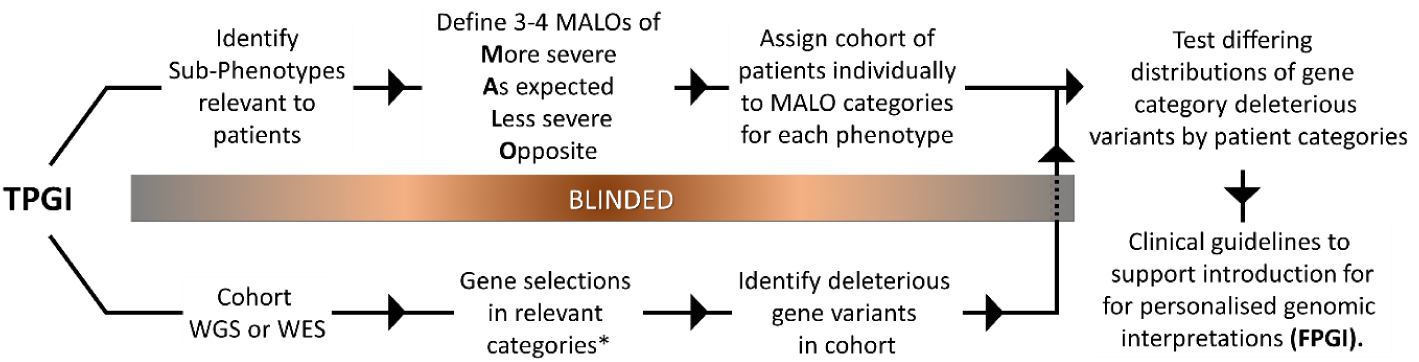
Framework for studies ‘towards personalised genomic interpretations’ (TPGI). The ‘MALO’ phenotypic severity scale refers to **M**ore severe than expected for disease; **A**s expected for disease; **L**ess severe than expected for disease, and (if feasible) **O**pposite phenotype. WGS, whole genome sequencing; WES, whole exome sequencing. *Gene selection by any broadly-relevant set of panels such as PanelApp established for 100,000 Genomes Project interpretations,[46] and is suggested to include categories not related to all phenotypes (as in current study) to control for methodological enrichments. Where TPGI demonstrates evidence of relevance across a patient cohort, this supports attention to a subsequent ‘for personalised genomic interpretations’ (**FPGI)** stage to be developed within mainstream medicine.

The hemorrhage and thrombosis phenotypes provided greater statistical power, since, as indicated in *Figure 3*, these can be considered to represent opposing ends of a spectrum. There were trends for deleterious variants in both coagulation genes (*Figure 4A upper middle pie charts*), and platelet genes (*Figure 4A upper right pie charts*) to be distributed towards the more severe hemorrhagic categories. Examining three-point scales of severe hemorrhage -usual hemorrhage – thrombosis, using non parametric Spearman’s correlations, deleterious (CADD>15) variants were enriched in patients with more severe hemorrhage, for both platelet (ρ = 0.25, p=0.008), and coagulation (ρ = 0.21, p=0.024) gene categories (*Figure 4B*).

## DISCUSSION

We have shown for 75 hematological genes where variants can cause heritable human disease, that deleterious variants are commonly found in man. Despite each variant individually being rare (allele frequency < 0.3%), overall more than a third of patients with an inherited vasculopathy that places them at risk of hemorrhage and anemia had at least one likely deleterious variant in diverse genes that could augment or modify these disease complications. In particular, patients with more severe hemorrhage that had been attributed solely to their diagnosed condition had a higher number of rare, deleterious variants in unrelated platelet and coagulation genes located across the genome. These findings provide evidence that even modestly-sized rare disease cohorts provide a framework for accelerating delivery of informed clinical care through whole genome sequencing using interpretative approaches based on phenotype-based targeting of genes with established pathogenic potential.

Most attention on the potential of DNA sequencing to personalise medicine focuses on discovery of novel genetic causes of disease,[1,43] and development of polygenic risk scores from large-scale studies identifying common variation across blocks of genes co-inherited in linkage disequilibrium[44] that can direct attention to novel *cis-*regulatory elements.[44,45] The presented approach offers an accelerated route to the clinic, by proceeding from genes already known to have functional impact in processes potentially relevant to the patient. While a small proportion of rare deleterious variants cause recognised human diseases, most do not, either because they are not present at sufficient zygosity, or because they are not in an appropriate patient/environment milieu to be exposed above an essentially compensated perturbation to normal physiology. Major databases emphasise the very high rates of rare deleterious variants in the human general population.[38] The current study offers an indicative example of how aberrant physiology imposed by an endothelial vasculopathy appears to augment the impact of variants in separate systems that might otherwise be clinically insignificant. More generally, the approach can be adopted in a framework of personalised genomic interpretations (PGIs) either in ‘towards PGI’ studies as in the current manuscript to provide statistical support, or ‘for PGI’ studies to deliver what is genuinely personalised, N=1 medicine (*Figure 5*).

Strengths of the study include WGS of a respectable number of rare disease patients with detailed phenotypic characterisation, spanning a range of severity phenotypes. Unusually for a hospital recruited-population, not all were referred because of personally severe features, with some presenting through family-based screening programmes. Nevertheless, a weakness of the study is that the group did have a bias towards a hospital presentation, and we cannot rule out that the burden of deleterious rare variants would be lower in a community-recruited population, potentially with milder phenotypic spectra. That said, the cohort were not recruited to the 100,000 Genomes Project because of a bleeding, anaemia, thrombotic, or infective phenotype, but instead because they met diagnostic criteria for a separate disease, the endothelial vascular dysplasia HHT. As such they provide an important window on the potential for similar scale, WGS-based high definition analyses in other diseases where phenotypes could be modified by concurrent, otherwise minor diatheses.

All variants identified were rare, at MAFs <0.5%, and therefore do not include two widely distributed polymorphic low expression alleles of *SPTA1. Figure 2Bii* indicates that with one exception, there was no reason for variants to segregate with the vasculopathy in families, since almost all variants were widely separated from the HHT genes, usually on different chromosomes. *AK1* is in linkage disequilibrium with *ENG*, and the single *AK1* deleterious variant was identified in an *ENG* family indicating the two are likely to co-segregate in that particular family. However, there was only one variant with a CADD score >15 that was identified in two members of the same family (an *ACVRL1* family). The findings could be extended by incorporating additional genes and categories, and in later individual-level considerations, it may be helpful to distinguish genes associated with dominant phenotypes (such as *ANK1* and *SPTB*) from others in which heterozygous variants are recessive. Further, it is not the place of the current manuscript to explore how these and the wider variants might change HHT clinical practice-such considerations with patient-level integrations are underway through the wider NHS Rare Disease Collaborative Network for HHT and Genomics England Clinical Interpretation Partnership (*Figure 5*). That said, some overview comments are appropriate:

- 11/104 HHT patients were heterozygous for a total of 11 deleterious variants (10 unique variants, 10 different families), in genes where deficiency is known to impact on platelet function leading to bleeding disorders. Furthermore, these variants were found more commonly in patients defined as having more severe hemorrhage either spontaneously, or precipitated for example by pregnancy, antiplatelet agents, anticoagulants or iron. Currently no attention in HHT management focusses on platelet disorders, yet the current study suggests such considerations alongside modifications to commonly prescribed drugs may be relevant to at least 10% of HHT patients.
- Similarly 13/104 HHT patients were heterozygous for a total of 14 deleterious variants (all unique variants, and in different families), in genes where deficiency is known to impact on the coagulation cascade. The data suggest this may be relevant to more than 12% of HHT patients.
- 24/104 HHT patients were heterozygous for a total of 32 rare deleterious variants (all unique) in genes where deficiency is associated with inherited hemolytic anemias, due to an increased rate of red cell destruction. Again, currently no attention in HHT anemia management focusses on extending the lifespan of endogenous erythrocytes as they suffer additional stresses on passage through HHT vasculature. Cross-over strategies with hemolytic anemias may be relevant to 1 in 4 HHT patients, with particularly attention directed to the subgroup of HHT patients currently receiving intense, intravenous iron regimes on weekly or near-weekly basis.[2,16]

The data suggest that there was no *a priori* reason why the study cohort had their rare, deleterious variants, with the platelet and coagulation deleterious gene variants enriched less compared to GnomAD 3.1, than other categories examined. Instead, comparison with population-level databases predict comparable burdens in other patient populations, drawing attention to new foci for research. For example, while genetic variants predisposing to pathogenic venous thromboses are usually assumed to encode components and mediators of the coagulation cascade, the current cohort suggested a possible trend with erythrocyte enzyme variants, and there has been some discussion of thrombosis in red cell enzymopathy fields.[47] Opportunities to test current associations will be enhanced if enzymopathy genes are included in gene panels for VTE, and further scientific examination appears warranted. Similarly, the observed enrichment of deleterious erythrocyte membrane gene variants in patients experiencing severe deep-seated infection warrants further study in the setting of the common (daily) bacteremias that can seed abscesses by currently unknown mechanisms.[19,48] Mechanistic examination of whether erythrocyte membrane changes modify macrophage opsonisation rates[49] may influence clinical risk assessments, resulting in extension or restriction of HHT patient cohorts for whom prophylactic antibiotics are recommended.[19-22] Notably, in both study cohort and GnomAD 3.1 populations, the greatest proportion of variants were for the red cell membrane category, where relevance to evolutionary fitness could be postulated.

We conclude that the potential to unmask pathophysiologically-relevant processes is augmented by supplementing binary phenotypic discriminators with severity scales that additionally influence risk-benefit considerations inherent in therapeutic provision. Accelerated pathways to more informed clinical trial design and personalised medical care can be the expected outcome of whole genome sequencing when analytics focus on rare deleterious variants for secondary clinical phenotypes that matter to patients and health care services.

## METHODS

### Clinical cohort recruitment

Patients attending the Hammersmith Hospital/West London Genomic Medicine Centre for management of HHT were recruited to the 100,000 Genomes Project if the family’s HHT pathogenic variant was not already known.[50,51] These comprised patients previously genotyped with no known causal gene identified, and a larger unselected group who had not undergone prior genetic testing and were offered the opportunity to participate during attendance at the clinical service during the recruitment period. Recruitment was offered to all patients attending on those days if they met the clinical diagnosis of HHT [52], and did not have a known genetic cause of HHT. The maximal number of patients was approached, and there was no bias in recruitment to patients with milder or more severe HHT features.

As described for the wider cohort,[7,16,20,53,54] all individuals reviewed in the clinical service were assessed using standardised methods including a detailed clinical history specifically focusing on aspects of the HHT phenotypes, and any disproportionate or unexpected symptomatic manifestations. Standardised hematological and biochemical assessments were performed using the hospital service laboratories as described.[7,16,20,53]

### Whole Genome Sequencing

The 100,000 Genomes Project was set up by the UK Department of Health and Social Security in 2013, to sequence whole genomes from National Health Service (NHS) patients recruited through one of 13 Genomic Medicine Centres (GMCs). Patient/family DNA sampling was accompanied by phenotypic data entries and integration with NHS health records. HHT was successfully nominated for inclusion in 2015. During data model set up, human phenotypic ontology (HPO) terms[55] were defined to capture the full range of phenotypes observed in HHT patients, totalling 301 different terms spanning disease manifestations, accessory and incidental phenotypes.[7,55] For each HHT patient recruited between September 2016 and August 2019, relevant HPO terms were submitted to Genomics England following patient recruitment using Open Clinica and Genie.[50]

Genomics England performed all whole genome sequencing (WGS) using the Illumina WGS Service Informatics pipeline for sequence alignments to the National Center for Biotechnology Information Genome Reference Consortium Human (GRCh) build 38/hg38,[39] and variant calling as described.[50] Anonymised outputs from Genomics England internal bioinformatics pipelines and analyses were examined in the Genomics England Research Environment through LabKey.[50]

### Candidate modifier gene selection and analysis

Genes were primarily selected based on causal influences on coagulation, hemorrhage, and/or red blood cell (erythrocyte) survival in the general population,[33,34] irrespective of whether a single deleterious allele would be disease-causing (dominant [heterozygous] or X-linked recessive [hemizygous]), or usually silent (e.g. autosomal recessive, such as thalassaemia or Sickle Cell traits]. The 75 selected genes were categorised by assigning to the single category for their most recognised role across hemoglobin production (i.e. ‘hemoglobinopathy’ genes), red cell (erythrocyte) membrane production (i.e ‘membranopathy’ genes), red cell enzymes (i.e. ‘enzymopathy’ genes); the coagulation cascade, and platelet biology (*Supplementary Table 1*).

Blinded to clinical sub-classifications described below, all DNA sequence variants in patients recruited from West London GMC under the category of HHT were identified in the selected hematological genes (*Supplementary Table 1*). Variant details (number, allele frequencies and CADD scores), and separately, categorised patient numbers, were approved for export through the Research Environment AirLock under subprojects RR42 (HHT-Gene-Stop), and RR43 (Genes, and AVM Development, Physiology, and Compensations in the Pulmonary Circulation (GADP-PAVMs). For export of categorised patient data, identifiers were converted to new codes to prevent inadvertent reveals to the clinical team who generated the phenotypic subcategories-for the purposes of ongoing studies, they remain blinded to patient-level genomic data for the hematological genes.

### Gene-level analysis

The burden of usual genetic variation was compared between genes and gene categories by two separate measures. The first used the gene damage index (GDI) which is a genome-wide, gene-level metric of the mutational damage that has accumulated in the general population, and performs well at removing exome variants in genes irrelevant to disease.[31] Genic intolerance was measured by the residual variation intolerance score (RVIS [32]), which performs better for detection of true positives, i.e. genes where newly-identified variants are more likely cause a recognised disease.

### Variant-level analysis

To evaluate distributions of affected genes and variants across the genome, the number of genes with variants, and number of variants were examined per Mb of DNA for each chromosome, and Circos[40] was used on the Galaxy platform[41] to generate a circular ideogram of GRCh38/hg38[36] for visualisations.

To rank likely deleteriousness of identified variants within the study cohort, Combined Annotation Depletion (CADD) scores were extracted from the University of Washington CADD server.[35-37] CADD scores rank the deleteriousness for all 9 billion single nucleotide variants, millions of small indels, and splice site variants based on machine learning trained on diverse genomic features derived from surrounding sequence context, gene model annotations, evolutionary constraint, epigenetic measurements and functional predictions.[35-37] A CADD score greater than 15 is commonly used as a single threshold to indicate likely deleteriousness,[35,42] and performed better than other single measures of deleteriousness using the mutation significance cutoff (MSC) gene-level thresholds for variant-level predictions.[42] With study methodology precluding integration within American College of Medical Genetics-Association for Molecular Pathology clinical care frameworks,[56,57] to enhance predictive power of variant pathogenicity calls, gene-level adjustments were incorporated. For these, we used the MSC 90% and 95% scores which define the 90% and 95% confidence interval lower boundaries of CADD scores of all high quality mutations described in that gene as pathogenic.[42] Generally, variants were categorised as CADD >15 (likely deleterious, incorporating all start loss, stop gain, frameshift, splice donor, splice acceptor, and a small proportion of missense and inframe deletion variants[57]); CADD 5-15 (uncertain significance, comprising most missense and inframe deletions, and some 5’ untranslated region and splice region variants); and CADD <5 (likely benign, comprising most synonymous, 5’ untranslated region, splice region and intronic variants). The 90% MSC [42] was used to identify genes where variants with lower CADD scores were more likely to be deleterious, and in the final analyses examining deleterious variants against patient phenotypic severity scales, comparisons were made with and without incorporation of gene-specific lower CADD score variants into the deleterious category.

The GnomAD 3.1.1 database, comprising 76,156 genomes and exomes was used as a comparator population.[38] For each of the 75 study genes, we extracted the total number of variants in GnomAD 3.1.1, and likely deleterious variants defined as those flagged as start loss, stop gain, frameshift, splice donor, splice acceptor, loss-of-function, and/or ClinVar-listing as likely pathogenic or pathogenic variants. In view of the differing approaches to classifications (molecular subtype/ClinVar assignments in GnomAD 3.1.1; CADD scores in study cohort), it was not appropriate to compare rates of deleterious variants by genes directly between GnomAD 3.1.1 and the study population. Instead, differential rates were compared between the categories of hematologic genes, enabling less relevant categories to control for methodological sources of enrichment. GnomAD 3.1.1 was also used to define population allele frequencies for the variants identified in the study cohort.

### Disease Sub-categorisations

Based on updated clinical information on the recruited patients by 28^th^ April 2021, continuous variables, and severity scales as currently used in our service [7,9,16,53,54] were assigned to all recruited patients, blinded to genotypes beyond the causal HHT variants emerging through clinical diagnostic pipelines:[51]

- HHT-related bleeding was categorised as H0 (as expected for HHT); H1 (precipitated excessive bleeds, i.e. severe bleeds precipitated by a pharmaceutical agent [10,27,28,30], or pregnancy-associated PAVM hemorrhage [14,58]); and H2 (spontaneous excessive bleeds). H2 was assigned for severe daily nosebleeds or gastrointestinal bleeds resulting in hemorrhage adjusted iron requirements (HAIR)[9] exceeding replacement feasibility using oral iron supplementation (resulting in intravenous iron and/or blood transfusion dependency),[16] or for a major spontaneous hemorrhage from a pulmonary[59] or cerebral[15] AVM.
- Anemia was categorised as A0 (within expected range for hemorrhagic losses and iron intake [9,16], and A1 (anemia out-of-proportion to the degree of hemorrhage based on HAIR[9] and the total iron intake from the diet and therapeutic oral or intravenous iron, as detailed in [16]).
- The venous thromboemboli (VTE) categories were T0 (no VTE), T1 (drug, peri-operative or otherwise precipitated VTE/pulmonary embolus), and T2 (spontaneous VTE/pulmonary embolus, using definitions as in [18]).
- Deep-seated infection categories were I0 or I1, with deep seated infection type listed for I1[19-22].

### Data Analysis

Summary and comparative statistics were generated using STATA IC v15.0 (Statacorp, College Station, US), STATA IC v16.0 (Statacorp, College Station, US), and GraphPad Prism 9 (GraphPad Software, San Diego, CA). One-way differences between categories were compared by Chi-squared test, and between continuous variables using Kruskal Wallis with Dunn’s multiple comparison test to derive p-values for pre-selected pair-wise comparisons. Two-way analyses to incorporate both data source and gene categories were performed using a two-way analysis of variance. The strength and direction of association between ranked variables were examined using Spearman’s rank-order correlation. Data were represented using graphics generated in GraphPad Prism 9 (GraphPad Software, San Diego, CA) and STATA IC v 16.0.(Statacorp, College Station, TX).

## Data Availability

Data Availability Statement
Primary data from the 100,000 Genomes Project, which are held in a secure Research Environment, are available to registered users. Please see https://www.genomicsengland.co.uk/about-gecip/for-gecip-members/data-and-data-access for further information.

## ACKNOWLEDGEMENTS

This research was made possible through access to the data and findings generated by the 100,000 Genomes Project. The 100,000 Genomes Project is managed by Genomics England Limited (a wholly owned company of the Department of Health and Social Care). The 100,000 Genomes Project is funded by the National Institute for Health Research (NIHR) and NHS England. The Wellcome Trust, Cancer Research UK and the Medical Research Council have also funded research infrastructure. The 100,000 Genomes Project uses data provided by patients and collected by the National Health Service as part of their care and support. We thank the National Health Service staff of the UK Genomic Medicine Centre and the families for their willing participation. This research was co-funded by the NIHR Imperial Biomedical Research Centre. The views expressed are those of the authors and not necessarily those of funders, the NHS, the NIHR, or the Department of Health and Social Care.

## DATA AVAILABILITY STATEMENT

Primary data from the 100,000 Genomes Project, which are held in a secure Research Environment, are available to registered users. Please see https://www.genomicsengland.co.uk/about-gecip/for-gecip-members/data-and-data-access for further information.

## AUTHOR CONTRIBUTIONS

KEJ co-developed the project design; selected gene categories and genes; performed all data analysis within the Genomics England Research Environment and GnomAD 3.1; exported anonymised datasets for numerical interrogation; retrieved all CADD scores; performed quantitative trait association studies, contributed content to Figures 1-4 (1C; 2B-D; 3C; 4B), and contributed to manuscript writing. EO and SB assisted with patient evaluations. FN, KO and KF recruited patients. MS and GT processed patient samples. GT, TF, JR and CLS contributed to project set up at West London Genomic Medicine Centre, and patient recruitment. CMM contributed to coagulation discussions. NC and DML contributed to red cell discussions. MJC set up, and FBP facilitated anonymised analyses within the Genomics England Research environment. GERC performed WGS, sequence alignments and variant calling. CLS set up the HHT-specific projects with Genomics England, the Respiratory Clinical Interpretation Partnership (GeCIP), and at West London GMC, reviewed the patients; categorised phenotypic severities; co-developed the project design; performed remaining data analyses, generated Figures 1-6, and wrote the manuscript. KEJ, CMM, NC, DML and CLS contributed to manuscript revisions. All authors reviewed and approved the final manuscript. CLS is responsible for the overall content as guarantor.

## DATA SUPPLEMENT

**Supplementary Table 1.** The 75 selected hematological genes by category.

| HGNC Symbol | Gene name | General population relevant phenotype (s) | GnomAD deleterious | Any variant | Total variants | CADD >15 variants |
| --- | --- | --- | --- | --- | --- | --- |
| **Haemoglobin production** |  |  |  |  |  |  |
| ATRX | ATRX chromatin remodeller | Alpha-thalassemia myelodysplasia syndrome | 75 | 1 | 1 | 0 |
| CDAN1 | Codanin 1 | Dyserythropoietic anemia, TYPE Ia; | 19 | 1 | 2 | 1 |
| HBA1 | Hemoglobin alpha | Alpha thalassemia | 29 | 0 |  |  |
| HBA2 | H Hemoglobin alpha emoglobin alpha |  | 51 | 0 |  |  |
| HBB | Hemoglobin beta | Beta thalassemia | 315 | 0 |  |  |
| HBG2 | Hemoglobin gamma | Fetal hemoglobin quantitative trait | 6 | 1 | 1 | 0 |
| HbS1L | HbS1-like translational GTPase | Beta thalassemia /Sickle cell disease/other | 0 | 1 | 1 | 1 |
| HBZ | Hemoglobin subunit zeta (embryonic) | Alpha thalassemia | 0 | 1 | 2 | 2 |
| KIF23 | Kinesin family member 23 | Dyserythropoietic Anemia, type III | 1 | 1 | 1 | 1 |
| KLF1 | Kruppel-like factor 1 | Dyserythropoietic Anemia, type IV | 15 | 0 |  |  |
| SEC23B | SC 23 homologue B | Dyserythropoietic Anemia, type II | 22 | 0 |  |  |
| **Erythrocyte membrane production** |  |  |  |  |  |  |
| ANK1 | Ankyrin | Hereditary spherocytosis type 1 | 37 | 1 | 2 | 2 |
| EPB41 | Erythrocyte Membrane Protein Band 4.1 | Hereditary elliptocytosis | 6 | 1 | 4 | 3 |
| EPB41L1 | Erythrocyte Membrane Protein Band 4.1 Like 1 | Not yet defined | 2 | 1 | 3 | 0 |
| EPB41L2 | Erythrocyte Membrane Protein Band 4 1 Like 2 | Hereditary elliptocytosis | 0 | 1 | 1 | 1 |
| EPB41L3 | Erythrocyte Membrane Protein Band 4.1 Like 3 | Not yet defined | 0 | 1 | 3 | 0 |
| EPB41L4A | Erythrocyte Membrane Protein Band 4.1 like 4A | Not yet defined | 1 | 0 |  |  |
| EPB41L4B | Erythrocyte Membrane Protein Band 4.1 like 4B | Giant hemangioma | 0 | 1 | 2 | 1 |
| EPB41L5 | Erythrocyte Membrane Protein Band 4.1 Like 5 | Not yet defined | 0 | 1 | 1 | 1 |
| EPB42 | Erythrocyte Membrane Protein Band 42 | Hereditary spherocytosis type 5 | 8 | 0 |  |  |
| KCNN4 | Potassium channel, calcium-activated, intermediate/small conductance, subfamily N, member 4 | Dehydrated hereditary stomatocytosis 2 | 4 | 1 | 2 | 0 |
| PIEZO1 | Piezo-type mechanosensitive ion channel component 1 | Dehydrated hereditary stomatocytosis | 38 | 1 | 24 | 3 |
| SLC4A1 | Solute carrier family 4 (anion exchanger), member 1; | Hereditary spherocytosis type 4 | 47 | 0 |  |  |
| SLC4A1AP | solute carrier family 4 adaptor protein | - | 0 | 1 | 2 | 0 |
| SPTA1 | Erythrocytic alpha spectrin | Hereditary elliptocytosis-2, pyropoikilocytosis, & spherocytosis, type 3 | 49 | 1 | 6 | 3 |
| *SPTB* | Erythrocytic beta spectrin | Hereditary spherocytosis type 2 | 27 | 1 | 6 | 5 |
| ***Erythrocyte enzymopathies*** |  |  |  |  |  |  |
| *AK1* | Adenylate kinase 1 | Nonspherocytic hemolytic anemia | 7 | 1 | 1 | 1 |
| *BPGM* | Bisphosphoglycerate Mutase | Hemolytic anemia | 5 | 1 | 1 | 1 |
| *ENO1* | Enolase 1 | Not yet defined | 0 | 0 |  |  |
| *G6PD* | glucose-6-phosphate dehydrogenase | Acute hemolysis; severe chronic non-spherocytic hemolytic anemia | 44 | 1 | 1 | 0 |
| *GCLC* | Glutamate-Cysteine Ligase Catalytic Subunit | Hemolytic anemia | 2 | 0 |  |  |
| *GPI* | Glucose-6-Phosphate Isomerase | Nonspherocytic hemolytic anemia | 11 | 1 | 1 | 1 |
| *GPX1* | Glutathione Peroxidase 1 | Hemolytic anemia | 0 | 1 | 1 | 1 |
| *GSR* | Glutathione-Disulfide Reductase | Hemolytic anemia | 2 | 0 |  |  |
| *GSS* | Glutathione synthetase | Hemolytic anemia | 10 | 1 | 2 | 1 |
| *HK1* | Hexokinase 1 | Hemolytic anemia | 9 | 1 | 1 | 1 |
| *NT5C3A* | Pyrimidine 5-prime-nucleotidase | Hemolytic anemia | 11 | 0 |  |  |
| *PGK1* | Phosphoglycerate kinase 1 | Hemolytic anemia | 16 | 1 | 1 | 1 |
| *PKLR* | Pyruvate Kinase | Chronic hereditary nonspherocytic hemolytic anemia | 24 | 0 |  |  |
| *TPI1* | Triosephosphate Isomerase 1 | Hemolytic anemia | 9 | 1 | 1 | 1 |
| ***Coagulation*** |  |  |  |  |  |  |
| *F2* | Thrombin (coagulation factor II) | Prothrombin deficiency/ thrombosis and dysprothrombinemia | 17 | 0 |  |  |
| *F2R* | Coagulation Factor II Thrombin Receptor | Thrombosis | 0 | 1 | 1 | 0 |
| *F2RL1* | Coagulation Factor II Thrombin Receptor-like 1 | Not yet defined | 0 | 0 |  |  |
| *F2RL2* | Coagulation Factor II Thrombin Receptor-like 2 | Think you need to include | 11 | 0 |  |  |
| *F2RL3* | Coagulation Factor II Thrombin Receptor-like 3 | Not yet defined | 0 | 1 | 1 | 1 |
| *F3* | Tissue factor (coagulation factor III) | Disseminated Intravascular Coagulation (COVID and HIV roles) | 0 | 0 |  |  |
| *F5* | Coagulation factor V | Autosomal recessive hemorrhagic diathesis or an autosomal dominant form of thrombophilia, which is known as activated protein C resistance. | 28 | 1 | 2 | 2 |
| *F7* | Coagulation factor VII | Factor VII deficiency | 31 | 1 | 1 | 1 |
| *F8* | Coagulation factor VIII | Hemophilia A | 338 | 1 | 3 | 1 |
| *F9* | Coagulation factor IX | Hemophilia B | 157 | 0 |  |  |
| *F10* | Coagulation factor X | Factor X deficiency | 12 | 0 |  |  |
| *F11* | Coagulation factor XI | Factor XI deficiency | 91 | 1 | 3 | 3 |
| *F12* | Coagulation factor XII | Hereditary angioedema type III | 6 | 0 |  |  |
| *F13A1* | Coagulation factor XIII A chain | Factor XIIIa deficiency | 23 | 1 | 1 | 1 |
| *F13B* | Coagulation factor XIII B chain | Factor XIIIb deficiency | 6 | 1 | 1 | 1 |
| *PROC* | Protein C, Inactivator of coagulation factors Va And VIIIa | Protein C deficiency (prothrombotic) | 58 | 0 |  |  |
| *PROS1* | Protein S | Protein S deficiency (prothrombotic) | 43 | 1 | 1 | 0 |
| *SERPINC1* | Antithrombin-III | Antithrombin-III deficiency which constitutes a strong risk factor for thrombosis. | 58 | 0 |  |  |
| *VWF* | Von Willebrand Factor | von Willebrand disease | 233 | 1 | 4 | 4 |
| **Platelets** |  |  |  |  |  |  |
| *ACTN1* | Actinin Alpha 1 | Bleeding Disorder, Platelet-Type, 15 | 24 | 0 |  |  |
| *ANKRD26* | Ankyrin Repeat Domain 26 | Autosomal dominant thrombocytopenia-2 | 1 | 1 | 1 | 1 |
| *DIAPH1* | Diaphanous Related Formin 1 | Thrombocytopenia (with deafness) | 22 | 1 | 1 | 1 |
| *ETV6* | ETS Variant Transcription Factor 6 | Bleeding Disorder, Platelet-Type | 13 | 0 |  |  |
| *GNE* | Glucosamine (UDP-N-Acetyl)-2-Epimerase/N-Acetylmannosamine Kinase | Thrombocytopenia (with myopathy) | 91 | 1 | 2 | 2 |
| *GP1BA* | Glycoprotein Ib platelet alpha subunit | Platelet-type von Willebrand disease; Macrothrombocytopenia | 23 | 1 | 2 | 1 |
| *GP1BB* | Glycoprotein Ib platelet beta subunit | Macrothrombocytopenia | 16 | 0 |  |  |
| *HPS1* | HPS1, biogenesis of lysosomal organelles complex 1 | Hermansky-Pudlak syndrome | 55 | 1 | 1 | 1 |
| *HPS6* | HPS6, biogenesis of lysosomal organelles complex 2 subunit 3 | Hermansky-Pudlak syndrome | 27 | 0 |  |  |
| *ITGA2B* | Platelet Glycoprotein IIb/Integrin Subunit Alpha 2b | Platelet-type bleeding disorders, which are characterized by a failure of platelet aggregation | 99 | 1 | 1 | 1 |
| *ITGB3* | Platelet Glycoprotein IIIa/Integrin Subunit Beta 3 | Bleeding Disorder, Platelet-Type, 16 | 70 | 0 |  |  |
| *MYH9* | Myosin Heavy Chain 9 | Macrothrombocytopenia | 39 | 1 | 1 | 1 |
| *RASGRP2* | RAS Guanyl Releasing Protein 2 | Bleeding disorder, platelet-type, 18 | 13 | 1 | 2 | 1 |
| *RUNX1* | Runt-related transcription factor 1 | Platelet disorder, familial, with associated myeloid malignancy | 77 | 0 |  |  |
| *TBXA2R* | Thromboxane A2 receptor | Bleeding disorder, platelet-type, 13 |  | 1 | 1 | 1 |
| *TUBB1* | Beta tubulin | Macrothrombocytopenia | 12 | 1 | 1 | 1 |

## Genomics England Research Consortium

### The Genomics England Research Consortium

John C. Ambrose^1^; Prabhu Arumugam^1^; Roel Bevers^1^; Marta Bleda^1^ ; Freya Boardman-Pretty^1,2^; Christopher R. Boustred^1^; Helen Brittain^1^; Mark J. Caulfield^1,2^; Georgia C. Chan^1^; Greg Elgar^1,2^; Tom Fowler^1^; Adam Giess^1^; Angela Hamblin^1^; Shirley Henderson^1,2^; Tim J. P. Hubbard^1^; Rob Jackson^1^; Louise J. Jones^1,2^; Dalia Kasperaviciute^1,2^; Melis Kayikci^1^; Athanasios Kousathanas ^1^; Lea Lahnstein^1^; Sarah E. A. Leigh^1^; Ivonne U. S. Leong^1^; Javier F. Lopez^1^; Fiona Maleady-Crowe^1^; Meriel McEntagart^1^; Federico Minneci^1^; Loukas Moutsianas^1,2^; Michael Mueller^1,2^; Nirupa Murugaesu^1^; Anna C. Need^1,2^; Peter O’Donovan^1^; Chris A. Odhams^1^; Christine Patch^1,2^; Mariana Buongermino Pereira^1^; Daniel Perez-Gil^1^; John Pullinger^1^; Tahrima Rahim^1^; Augusto Rendon^1^; Tim Rogers^1^; Kevin Savage^1^; Kushmita Sawant^1^; Richard H. Scott^1^; Afshan Siddiq^1^; Alexander Sieghart^1^; Samuel C. Smith ^1^; Alona Sosinsky^1,2^; Alexander Stuckey^1^; Mélanie Tanguy^1^; Ana Lisa Taylor Tavares^1^; Ellen R. A. Thomas^1,2^; Simon R. Thompson^1^; Arianna Tucci^1,2^; Matthew J. Welland^1^; Eleanor Williams^1^; Katarzyna Witkowska^1,2^; Suzanne M. Wood^1,2^.

1. Genomics England, London, UK
2. William Harvey Research Institute, Queen Mary University of London, London, EC1M 6BQ, UK.

